# Delayed Start of the Respiratory Syncytial Virus Epidemic at the End of the 20/21 Northern Hemisphere Winter Season, Lyon, France

**DOI:** 10.1101/2021.03.12.21253446

**Authors:** Jean-sebastien Casalegno, Etienne Javouhey, Dominique Ploin, Martine Valette, Remi Fanget, Sandrine Couray Targe, Anne-Florence Myar-Dury, Muriel Doret-Dion, Mona Massoud, Phillipe Vanhems, Olivier Claris, Marine Butin, Florence Ader, Sylvie Bin, Alexandre Gaymard, Bruno Lina, Florence Morfin, VRS study group in Lyon, Yves Gillet

## Abstract

The implementation of Non Pharmaceutical Interventions (NPI), triggered by the emergence of covid-19, decrease the RSV circulation. Data, from our ongoing surveillance; show a late introduction of RSV at the end of December and a 4 month delayed epidemic start without significant change in our NPI policy. This data indicates that RSV still have the potential to give a late season outbreak in northern hemisphere. RSV surveillance should be reinforced and RSV Pharmaceutical Interventions maintained for at risk neonate

---

The emergence of COVID-19 triggered the massive implementation of Non-Pharmaceutical Interventions (NPI) including social distancing, school closures, travel restrictions, and the use of masks in public spaces. It is now certain that these policy measures have reduced the transmission of SARS-CoV-2 but have also affected other circulating respiratory viruses such as Respiratory Syncytial Virus (RSV) and Influenza Virus (1). Indeed, no active circulation of RSV or Flu was observed at the end of January 2021 in the northern hemisphere although initial incidence cases in 2019 and 2020 were observed as soon as september (2). Recent reports from Australia (3) described an inter-seasonal RSV epidemic in Australian children following the reduction of COVID-19–related public health measures from September 2020 to January 2021. This unexpected inter-seasonal RSV resurgence was also reported in South Africa (4).

As previously described, we prospectively collected hospital laboratory data (5) and newborn cohort data (6) from the University Hospital of Lyon (HCL; Hospices Civils de Lyon) for the purpose of RSV surveillance in the Lyon metropolitan area (approximately 1.5 million inhabitants). A case was defined as any laboratory-confirmed RSV detected by nucleic acid amplification. A severe case was defined as an admission in the first year of life to one of the conventional paediatric hospital departments with a RSV positive sample. The incidence rate of severe cases was estimated per 1,000 births in the HCL newborn cohorts (8 958 births in 2020).

The RSV epidemics have had a major annual seasonal pattern in Lyon with the first cases usually detected at week 41 followed by a peak at the end of the year around week 51 (6). In 2020, the first RSV cases of the 20/21 season were detected in Lyon at week 46 and 47 (Figure 1) at the same time of the southern hemisphere outbreak. A sustained detection of cases was observed from week 51, which is the expected time of the epidemic peak, to week 5. On week 6, the RSV epidemic was declared in the first French region (Ile de France) while the number of RSV cases has continued to increase in the Lyon population (7). Subtyping indicated a predominant RSV-A epidemic (50 RSV-A/52 RSV subtyped by week 9). A late peak in RSV circulation had already been observed in February in our population during the 2010/2011 RSV season. The reason for such late circulation remain unknown so far. The fact that it was the first season after the A(H1N1)pdm09 pandemic or the extreme climatic event observed during December 2010 (one of the colder december ever record) may have play a role. Taken together, these 2 atypical seasons highlight the potential for late RSV transmission when collective immunity is not reached.

**Figure 1:**
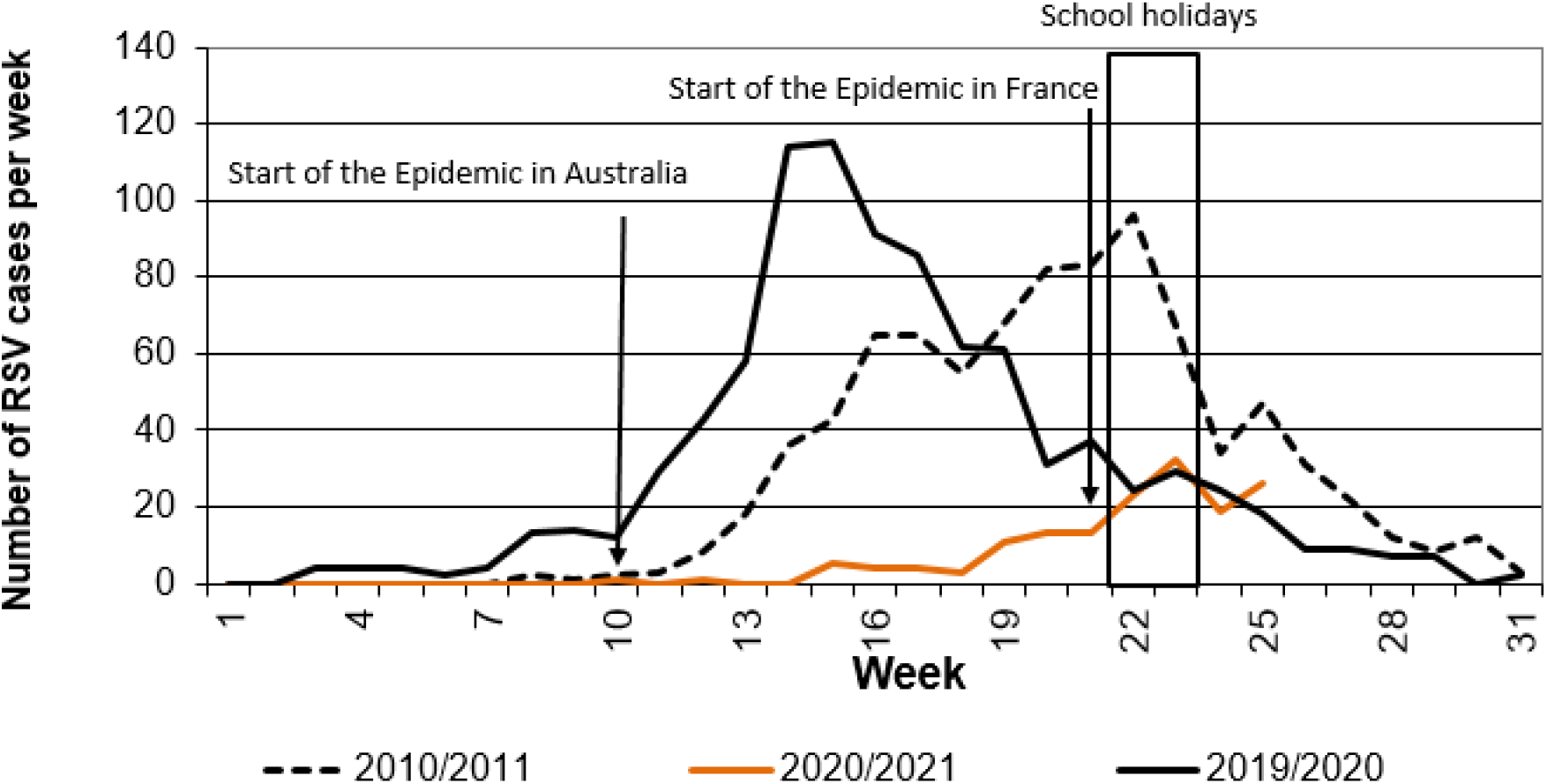
Number of RSV cases per week at the Lyon hospital virology department during the 2020/21, 2019/20, 2010/11 winter. Our surveillance data indicates that the first RSV were introduced in Lyon when an inter-seasonal RSV epidemic start in the southern hemisphere with a 8-month delay. The RSV epidemic is finally starting in Lyon with a 4-month delay at the end of the winter season without a significant change in the NPI strategy.

No obvious change in COVID-19 NPI measures since September 2020 could explain the increased number of RSV cases in our community by the end of February 2021. For example, the Oxford calculated stringency level for France varied from 63.89 in January to 72.22 end of February (8). The 2-week school closure due to spring holidays in our region (Weeks 7 to 8) may have delayed the start of the RSV epidemic with a significant increase in the number of cases observed on week 9.

At the time of this report (08/03/2021), the incidence rate for severe cases is still low in the 2020 birth cohort (4.8 (95% CI 4.0–7.0) severe cases per 1,000 live births) compared with the 2019 birth cohort (16.6 (95% CI 14.0–19.0) severe cases per 1,000 live births). As already reported, the incidence rate for severe cases is higher in premature newborns, defined as <37 weeks of gestational age (GA), (9.1 (95% CI 0.4–1.9) severe cases per 1,000 live births (6). Usually the incidence rate of severe cases is higher for the infant born before the peak (October and November) compared with those born after (January and February) (6) (Figure 2). For this late RSV season, we already observe a higher incidence rate of severe cases for the infant born in December 2020 and January 2021 (Figure 2).

**Figure 2:**
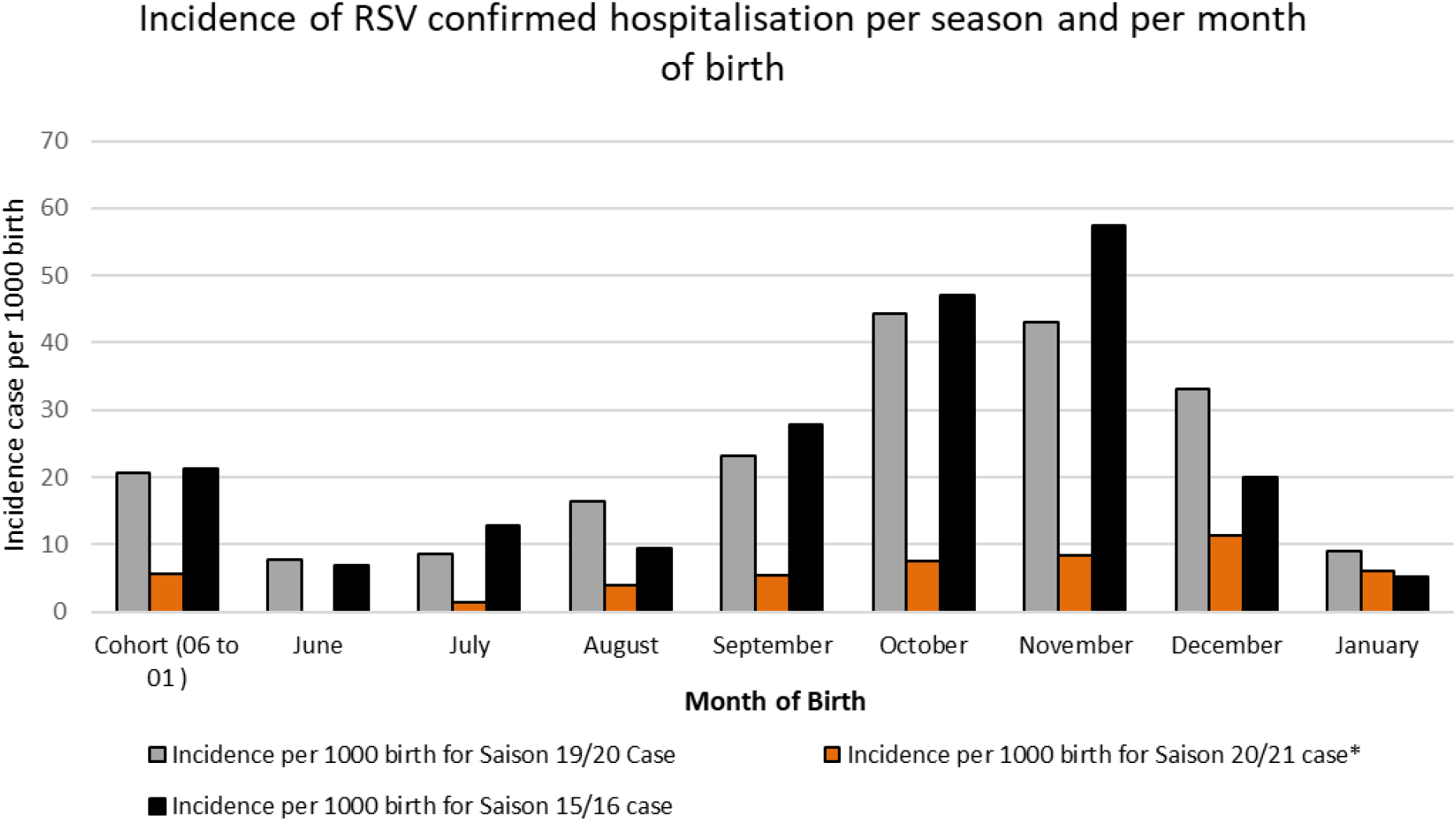
Incidence rate for RSV severe cases from three different birth cohorts (06-2015 to 01-2016; 06-2019 to 01-2020; 06-2020 to 01-2021) exposed to three RSV season (15/16, 19/20, 20/21). Incidence of hospitalisation with confirmed RSV infection the first year of life (severe case) is calculated per 1000 births and stratified per month of birth. *Provisional data, estimated at the time of the report (08/03/2021).

This observation highlights the risk of a late RSV outbreak for northern hemisphere cities or countries experiencing low levels of RSV circulation despite the implementation of COVID-19 NPI measures. It can be hypothesized that the seasonal transmission drivers combined with the increased susceptibility of the infant cohort and the waning RSV immunity in adults have the potential to overcome the NPI reduction in RSV transmission. NPI may still contribute to greatly reduce the RSV incidence in an at-risk population (infant and elderly), but it is too soon to measure its impact. Combined efforts of modelling, genome sequencing, and sero-epidemiological testing could shed an unprecedented light on the global RSV dynamic and the NPI impact. This is a major issue as the delayed circulation of RSV and Influenza in an increasingly susceptible population may have uncertain consequences on our public health system (9). Northern hemisphere countries should be aware of this probability and ensure that RSV pharmacological prevention is maintained for at-risk neonates and that RSV and Influenza surveillance is reinforced.

## Data Availability

Update Surveillance data are available on request

## Author’s Contribution

EJ, DP, MV, SCT, AFMD, MM, MDD, PV, BL, FM, VRS study group in Lyon, OC, MB, AF, YG, VRS study group performed the survey. JSC analysis the data and wrote the first draft. EJ, DP, MV, SCT, AFMD, PV, BL, FM, SB corrected the first draft.

## VRS study group in Lyon

Jean-sebastien Casalegno, Etienne Javouhey, Dominique Ploin, Martine Valette, Remi Fanget, Sandrine Couray Targe, Anne-Florence Myar-Dury, Muriel Doret-Dion, Mona Massoud, Phillipe Vanhems, Olivier Claris, Marine Butin, Florence Ader, Sylvie Bin, Alexandre Gaymard, Bruno Lina, Florence Morfin, VRS study group in Lyon, Yves Gillet4Antoine Ouziel, Jean-claude Tardy, Pascal Gaucherand, Jerome Massardier, Stephanie Polazzi, Antoine Duclos, Mehdi Benchaib, Regine Cartier, Marine Jourdain, Michelle Ottmann, Rolf Kramer, Sylvie Fiorini, Nathalie Rivat, Yahia Mekki, Julie Fort-Jacquier, Maud-Catherine Barral, Vey Noelie, Julie Haesebaert, Come Horvat, Leo Vidoni, Jean-Marc Reynes, Jean-Francois Eleouet, Laurence Josset, Matthieu Receveur, Gregory Queromes.

## Acknowledgements

We acknowledge the contribution of the pediatric department, laboratory technical staff and the BEHcl Team.

